# Mapping myelin alterations in Neurofibromatosis Type 1 using Magnetization Transfer and T1W/T2W Ratio Imaging

**DOI:** 10.64898/2025.12.09.25341878

**Authors:** Varun Arunachalam Chandran, Caroline Lea-Carnall, Yuping Yang, Anna Wild, Matthew McCowen, Grace Vassallo, Jonathan Green, Hamied Haroon, William Lloyd, Stavros Stivaros, Nils Muhlert, Shruti Garg

## Abstract

**Background:** Aberrant myelination represents a critical but understudied mechanism in cognitive difficulties associated with neurodevelopmental conditions. Neurofibromatosis 1 (NF1), provides a unique monogenic model to investigate this relationship, as white matter abnormalities are consistently observed, yet their microstructural basis remains uncharacterized. We present the first dual-modality quantitative myelin mapping study in NF1, employing Magnetization Transfer and T1W/T2W ratio imaging to delineate regional myelin alterations.

**Methods and materials:** We conducted a case-control study of 78 children (58 NF1 and 20 neurotypical controls, ages 11-18 years). Magnetization Transfer Ratio (MTR) and T1-weighted/T2-weighted (T1W/T2W) ratio imaging were used to quantify regional myelin properties. Working memory was assessed using the visuospatial n-back task.

**Results:** Compared to controls, children with NF1 showed significant myelin reductions in the thalamus, basal ganglia and cerebellum, converging across both imaging modalities. These deficits persisted independent of T2 hyperintensities, indicating a primary myelin pathology. Regional myelin alterations did not correlate with working memory performance.

**Conclusion:** This is the first study to employ two simultaneous methods to characterise myelin differences in NF1. The findings indicate that NF1-related myelin deficits are predominantly confined to the brainstem, cerebellar, and diencephalic structures. These results provide a neurobiological framework for understanding white matter pathology in NF1 and may help guide therapeutic strategies targeting myelination deficits in this disorder.

## Introduction

Myelin abnormalities represent a convergent pathophysiological mechanism across major neuropsychiatric conditions, including autism, Attention Deficit Hyperactivity Disorder (ADHD), schizophrenia and major depression (Haroutunian et al., 2014, Nave & Ehrenreich, 2014). Genetic variants affecting oligodendrocyte function and myelination contribute substantially to psychiatric risk, yet the neurobiological pathways linking myelin disruption to cognitive dysfunction remain incompletely understood (Alex et al., 2023, Douet et al., 2014). Myelination, which begins during the second trimester and extends through adolescence (Jakovcevski et al., 2009), is essential for neural circuit maturation underlying executive functions, including working memory, attentional control and decision-making (Corrigan et al., 2021, O’Muircheartaigh et al., 2014, Deoni et al., 2015, Yeung et al., 2014). Disrupted myelination, therefore, represents a plausible biological substrate for cognitive impairments across diverse clinical phenotypes.

Neurofibromatosis 1 (NF1) provides a genetically defined model for investigating how alterations in myelination contribute to cognitive difficulties and neural differences relevant to common neurodevelopmental conditions. This autosomal dominant disorder, which affects 1 in 3000 people (Lee et al., 2023), is caused by loss-of-function mutations in the *NF1* gene, a negative regulator of the Ras/MAPK and downstream signalling pathways. Cognitive and behavioural difficulties affect 70% of children with NF1 (Hachon et al., 2011), with elevated rates of co-occurring Autism Spectrum Conditions (ASC) in 10-30%, and ADHD in 50% of all cases. NF1 haploinsufficiency impairs oligodendrocyte maturation and myelin integrity (Asleh et al., 2020, Karlsgodt et al., 2012), which in turn are thought to contribute to learning impairments seen in the condition. Here, we employ converging multi-modal MRI methods to examine whether there are regional changes in markers of myelination in children with NF1.

Prior neuroimaging studies have yielded inconsistent findings regarding myelination in NF1. A recent semi-quantitative myelin imaging study using T1W/T2W ratio showed lower grey matter myelin content in widespread cortical regions and subcortical regions including accumbens in NF1 compared to typically developing controls (Plank et al., 2025^a^). Paradoxically, a quantitative T1-mapping technique showed increased white-matter myelination in various cortical brain regions and white matter fibre tracts in NF1 compared with controls (Plank et al., 2025^b^). Studies using diffusion tensor imaging show aberrant white matter microstructure in the major fibre tracts, including anterior thalamic radiation, inferior fronto-occipital fasciculus and corpus callosum suggesting myelination differences and/or changes in density of axonal packing (Siqueiros-Sanchez et al., 2024, Ferraz-Filho et al., 2012). Reduced fractional anisotropy and increased mean diffusivity in the anterior thalamic radiation, cingulum bundle and superior longitudinal fasciculus have been linked to executive function difficulties in NF1 (Koini et al, 2017). These discrepancies likely reflect methodological differences in myelin sensitivity and quantification approaches.

Here, we address these inconsistencies through multi-modal myelin imaging techniques in children and adolescents with NF1. We combined Magnetization Transfer Ratio (MTR)-a quantitative technique extensively validated in demyelinating disorders (Moccia et al., 2020, Chen et al., 2005), but not previously applied to NF1, with T1W/T2W ratio mapping. This multi-modal approach provides convergent evidence for regional myelin alterations and tests whether different imaging contrasts yield consistent findings. One advantage of using T1W/T2W ratios is that they are routinely acquired clinical MRI sequences, making them widely accessible. We hypothesized that the MTR and T1W/T2W ratios in grey and white matter would be lower in NF1 than in neurotypical controls. Further, we investigated the relationship between myelin and working memory performance and predicted that lower myelination in NF1 would be related to worse performance on executive function tasks.

## Methods and materials

Fifty-eight children and adolescents (Table 1) aged 11-18 years with NF1 were recruited from the Manchester Centre for Genomic Medicine NF1 clinics and through NF charities newsletters and social media pages. Eligibility criteria included (i) clinical diagnosis made using the National Institute of Health diagnostic criteria and/or molecular diagnosis of NF1 (Legius et al., 2021); (ii) no history of brain injury/intracranial pathology other than asymptomatic optic pathway or other asymptomatic and untreated NF1-associated white matter lesion or glioma; (iii) no history of epilepsy or any major mental illness and (iv) no MRI contraindications. Twenty neurotypical controls aged 11-18 years were recruited from the community via advertisements placed in the staff newsletters. Eligibility criteria for the controls included no preexisting neurodevelopmental or medical conditions. Written informed consent was obtained from the parents of all children with NF1 recruited to this study. The study was initiated after receiving approval from the NHS Research Ethics Committee (reference number: 18/NW/0762).

**Table 1.** Participant characteristics. *NT-Neurotypical Controls, VABS-ABC-Vineland Adaptive Behaviour Composite*.

| Variables | MTR sample |  | T1W/T2W sample |  |  |  |
| --- | --- | --- | --- | --- | --- | --- |
|  | NF1 (N=26) |  | NF1 (N=55) |  | NT (N=20) |  |
|  | Mean | SD | Mean | SD | Mean | SD |
| Age | 13.92 | 1.896 | 14.47 | 1.971 | 15.30 | 2.055 |
| Gender (M/F) | 15/11 |  | 27/28 |  | 8/12 |  |
| Pituitary depth | 6.197 | 1.133 | 5.801 | 1.049 | 6.456 | 0.701 |
| VABS-ABC | 77.72 | 16.759 | 72.54 | 18.509 | - | - |

### Behavioural assessments

The visuospatial n-back task was used to evaluate working memory performance in all NF1 participants. The n-back working memory task was designed using the E-prime (version 3.0, Psychology Software Tools) stimuli presentation software. These n-back tasks were performed at four different levels, including 0-back, 1-back, 2-back, and 3-back, with the task difficulty increasing at each stage. The visuospatial n-back task used blue squares shown on a black grid (3x3), except for the centre position. Participants were instructed to press the ‘A’ key, whenever they saw the target that matched the corresponding squares in accordance with various levels of complexity. We used all four levels of accuracy and response times to compare low and high working memory load performance with myelination. Accuracy was measured as the proportion of correct hits and correct misses corresponding to each trial within a block (averaged across each n-back condition), while the response time was measured as the time elapsed from the start of each trial to pressing the button. Principal Component Analysis with varimax rotation was applied to the visuospatial n-back working memory performance variables, including 0-back, 1-back, 2-back and 3-back accuracy and reaction times. Two components were retained, based on the eigenvalue-one criterion. For the MTR cohort, factors 1 and 2 associated most strongly with accuracy and reaction time respectively, whereas for the T1W/T2W cohort, factor 1 and 2 associated most strongly with reaction time and accuracy respectively factor 2 with (supplementary section 1: Table 5).

Vineland Adaptive Behavioural Scale (VABS-III), a parent-rated questionnaire, was administered to assess the communication, daily living skills, socialisation and adaptive behaviour composite scores (Carter et al., 1998).

### Magnetization Transfer Imaging and T1W/T2W ratio

Magnetization Transfer Imaging (MTI) is a myelin-sensitive quantitative MRI imaging technique that exploits interactions between the bound pool of protons (associated with myelin macromolecules) and the free water pool (mobile protons) (Wolff & Balaban 1989), Henkelman et al., 2001). When an off-resonance MT saturation pulse is applied, magnetization of the bound pool is selectively saturated. This saturation is then transferred to the free pool through cross-relaxation and chemical exchange, reducing the free water signal. By comparing images with and without the MT pulse, the Magnetization Transfer Ratio (MTR) images are derived, which is sensitive to myelin content and useful in studying demyelinating disorders particularly in multiple sclerosis. T1W/T2W ratio is another quantitative myelin mapping technique used to measure the myelin concentration and tissue microstructure (Glasser & Van Essen, 2011, Uddin et al., 2019). T1W/T2W ratio provides an enhanced contrast-to-noise ratio in measuring both grey and white matter myelin. This myelin mapping technique is comparable to imaging methods like myelin water fraction and magnetization transfer imaging (Pareto et al., 2020, Ganzetti et al., 2014). Unlike other quantitative myelin imaging techniques, T1W/T2W myelin mapping approach has the advantage of using conventional T1- and T2-weighted structural MRI brain images acquired for routine clinical practice and takes less time without having to apply advanced imaging protocols.

### MRI data collection

MRI data was acquired using a Philips Achieva 3T MRI scanner (Philips Healthcare, Eindhoven, Netherlands) with a 32-channel head coil. The structural MRI acquisition protocol included are: T1-weighted structural (on sagittal-plane): 3D fast field echo with repetition time (TR) = 8.40 ms, echo time (TE) = 3.84 ms, flip angle (FA) = 8°, voxel size = 0.94 x 0.94 x 1 mm^3^, matrix = 256 x 256, slices = 160. T2-weighted structural (on axial-plane): Turbo Spin echo with TR = 3756 ms, TE = 88 ms, FA = 90°, voxel size = 0.45 x 0.45 x 4 mm^3^, matrix= 512 x 512, slices = 40. Magnetization Transfer (MT) images (on axial-plane) were acquired with a 3D fast field echo, TR = 34 ms, TE = 3.4 ms, voxel size = 0.44 x 0.44 x 3 mm^3^, matrix = 512 x 512, number of slices = 45, FA = 12°. MT data was acquired with (MT_on_) and without (MT_off_) applying RF saturation pulses. T1W and T2W images were acquired from 58 subjects, whilst MTR images were acquired from 48 subjects. Three MRI datasets were excluded from the data analysis after quality control assessment and visual inspection. One structural MRI dataset demonstrated severe cortical atrophy and large periventricular dilation, whereas another two MTR datasets had excessive subject motion artefacts. After data quality assessment, MTR images were available for forty-eight participants (26 NF1, 20 controls) and T1W/T2W images were available for seventy-five participants (55 NF1, 20 controls) for analysis.

### MRI data processing

MRI data were preprocessed using the FSL analysis suite (FMRIB Software Library, Release 6.0 (c) 2018, The University of Oxford, https://fsl.fmrib.ox.ac.uk/fsl/fslwiki/FSL). T1W and T2W structural brain images were subjected to skull-stripping using FSL’s BET and bias field correction using fsl_anat pipeline. The T2W images were resampled to match the inplane voxel resolution of the T1W data and were then coregistered to the T1W images using FSL’s FLIRT (Jenkinson, M., & Smith, S. 2001). The T1-weighted images were segmented into grey matter (GM) and white matter (WM) partial volume maps. The grey and white matter tissue probability maps were subjected to 50% threshold to generate binary masks. The coregistered T1W and T2W data was used to produce T1W/T2W ratio maps and the segmented GM and WM brain images were used as masks to produce GM- and WM-specific T1W/T2W maps.

The MT data was skull-stripped (brain extraction tool) using FSL’s BET followed by corregistration of MT_on_ to MT_off_ using FSL’s FLIRT. The formula MTR=(MT_off_ – MT_on_)/MT_off_ x 100 was applied to the data using fsl_maths to generate an MTR map. The whole brain MTR map was coregistered to the corresponding subject’s T1W data using FLIRT. The T1W data was spatially normalized to the MNI152 standard space using fsl_fnirt and the associated transform was applied to the T1W/T2W, and MTR maps. The MTR and T1W/T2W (GM and WM maps) were smoothed to improve the signal to noise ratio within the local neighbourhood voxels using a Gaussian kernel (FWHM = 8 mm) and (FWHM = 2 mm) respectively.

Pituitary depth was used as a proxy measure to control for the effects of puberty (Sari et al., 2014). Pituitary depth was measured using the T1-Weighted structural MRI brain images from a sagittal plane, with clear visibility of the cerebral aqueduct, with the application of RadiAnt DICOM viewer https://www.radiantviewer.com/. Subsequently, a scale was placed covering the superior and inferior end to measure the depth of the pituitary gland. A General Linear Model was used to investigate associations between imaging measures (including MTR and T1W/T2W ratio maps) by applying permutation (n=5000) based testing. Specifically, we tested for case-control myelin differences between NF1 and controls with age, gender and pituitary depth as covariates. We tested the relationship between myelin (grey and white matter) and working memory (interaction effects) between NF1 and controls. In addition, we also tested the relationship between myelin (grey and white matter) and adaptive behaviour in NF1.

## Results

### Sample characteristics

Across the whole cohort (N=There were no significant group differences in age (p<0.058) or gender (p<0.485) between NF1 and controls (Table 1). The pituitary depth (p<0.006) was significantly (p<0.006) lower for NF1 compared to controls.

### MTR study

The study sample used for MTR (NF1= 26, controls= 20) showed a significant group difference in age (p<0.012). No significant group differences (Chi-square) were found in gender (p<0.234).

We used MTR as an exploratory study to examine the myelin differences in NF1. The MTR results showed significantly reduced grey matter myelin in the subcortical structures including the left thalamus and extended clusters to the left hippocampus in NF1 compared to controls (Figures. 1 and 2, Table 2). We also found a large cluster centered in the left pallidum, thalamus, extending to the brainstem and cerebellum in NF1 relative to controls. No significant association between myelin measured by MTR was found with working memory performance (accuracy or reaction time) or with adaptive behaviour (See supplementary section: 3).

**Fig. 1.**
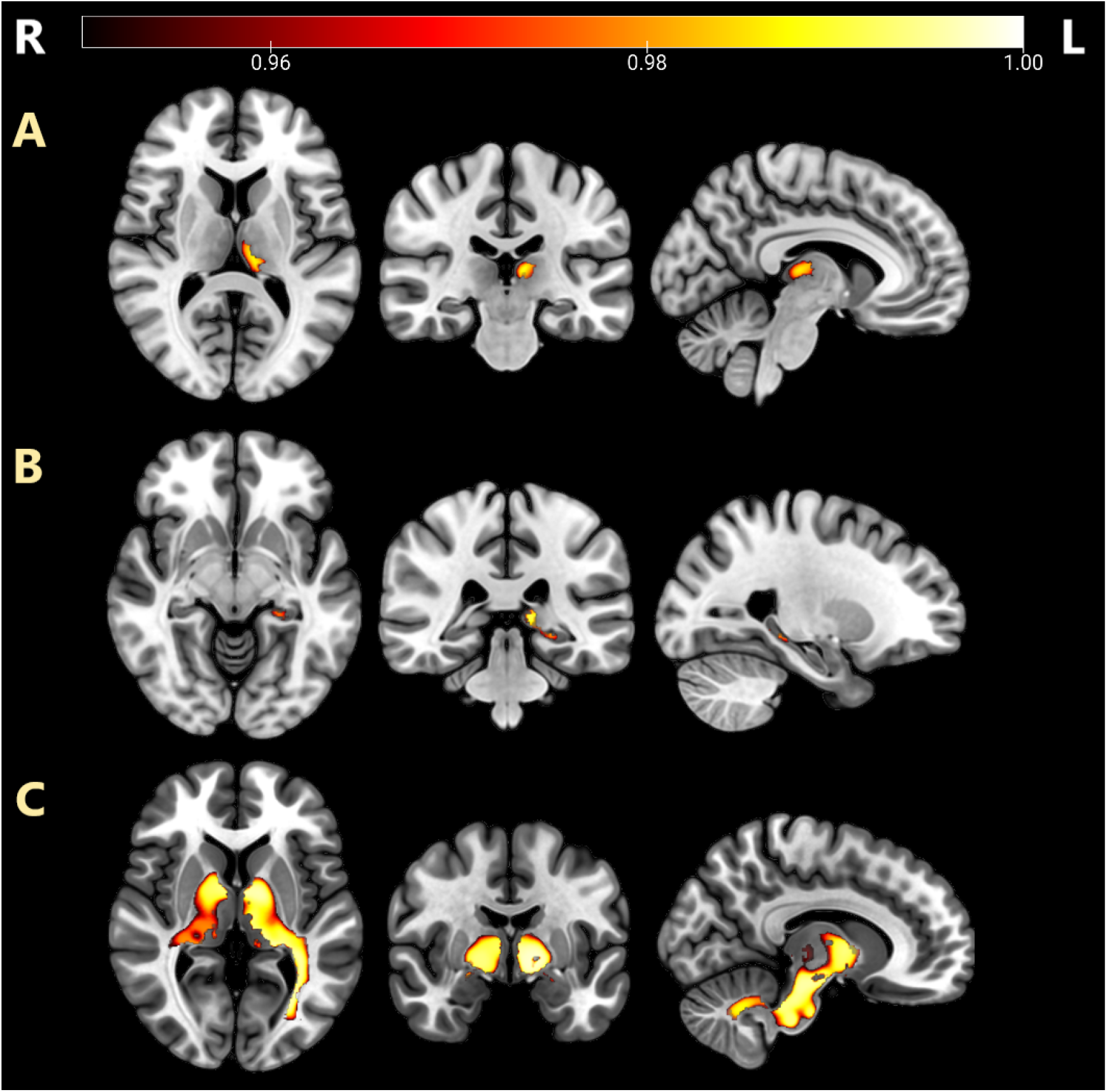
MTR clusters (on MNI152 brain template overlay) demonstrating regions of significantly lower myelin (red-yellow) in NF1 compared to controls. Grey matter myelin shows significant clusters in (A) left thalamus extended to (B) left hippocampus. (C) A large extended cluster of reduced white matter myelin is seen in thalamus, basal ganglia and cerebellum. A threshold (p=0.05) was used to delineate all clusters from family wise error corrected maps.

**Fig. 2.**
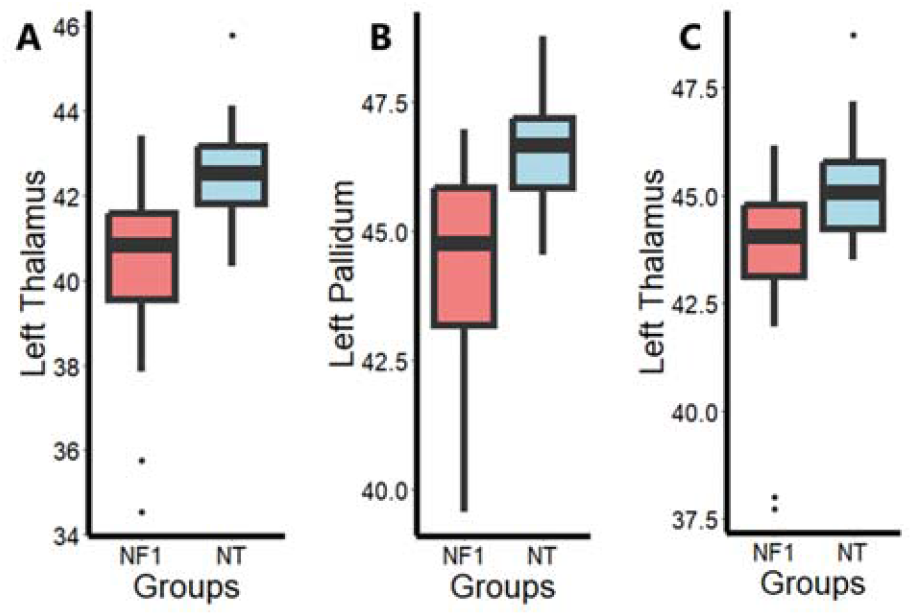
The boxplots for MTR represents the lower myelin in NF1 (red) than the controls (blue) in the grey matter including (A) left thalamus and white matter including (B) left pallidum and (c) left thalamus. The x-axis represents the groups and the y-axis represents the myelin. The clusters were extracted with a spherical ROI (radius= 2mm) for subcortical structures to generate all boxplots.

**Table 2.** Case-control comparison of MTR grey and white matter myelin between NF1 and controls *k-Cluster size, F-effect size, p<0.05* (Family Wise Error corrected).

| Brain regions | Hemisphere | MNI-coordinates |  |  | k | F | p-FWE |
| --- | --- | --- | --- | --- | --- | --- | --- |
| Grey matter myelin |  | x | y | z |  |  |  |
| Thalamus | Left | -13 | -35 | 4 | 1911 | 3.65 | 0.009 |
| White matter myelin |  |  |  |  |  |  |  |
| Pallidum | Left | -12 | -7 | -8 | 67127 | 2.96 | 0.003 |
| Thalamus | Right | -16 | -26 | 4 | 67127 | 3.80 | 0.050 |

### Sensitivity analyses

We repeated the MTR analyses for the 8 NF1 participants without T2WMH and compared them to controls. We found reduced white matter myelin in a large cluster originating from the left thalamus (x= -6, y= -3, z= 1; k= 1254, F= 4.84, p= 0.039, FWE corrected) extending across the left pallidum and midbrain in NF1 compared to controls (Fig. 3 and 4). We also found reduced grey matter myelin in the left thalamus (x= -17, y= -28, z= 4; k= 9474, F= 6.41, p= 0.001, uncorrected) extended to left hippocampus, however this cluster did not survive the multiple comparison correction.

**Fig. 3.**
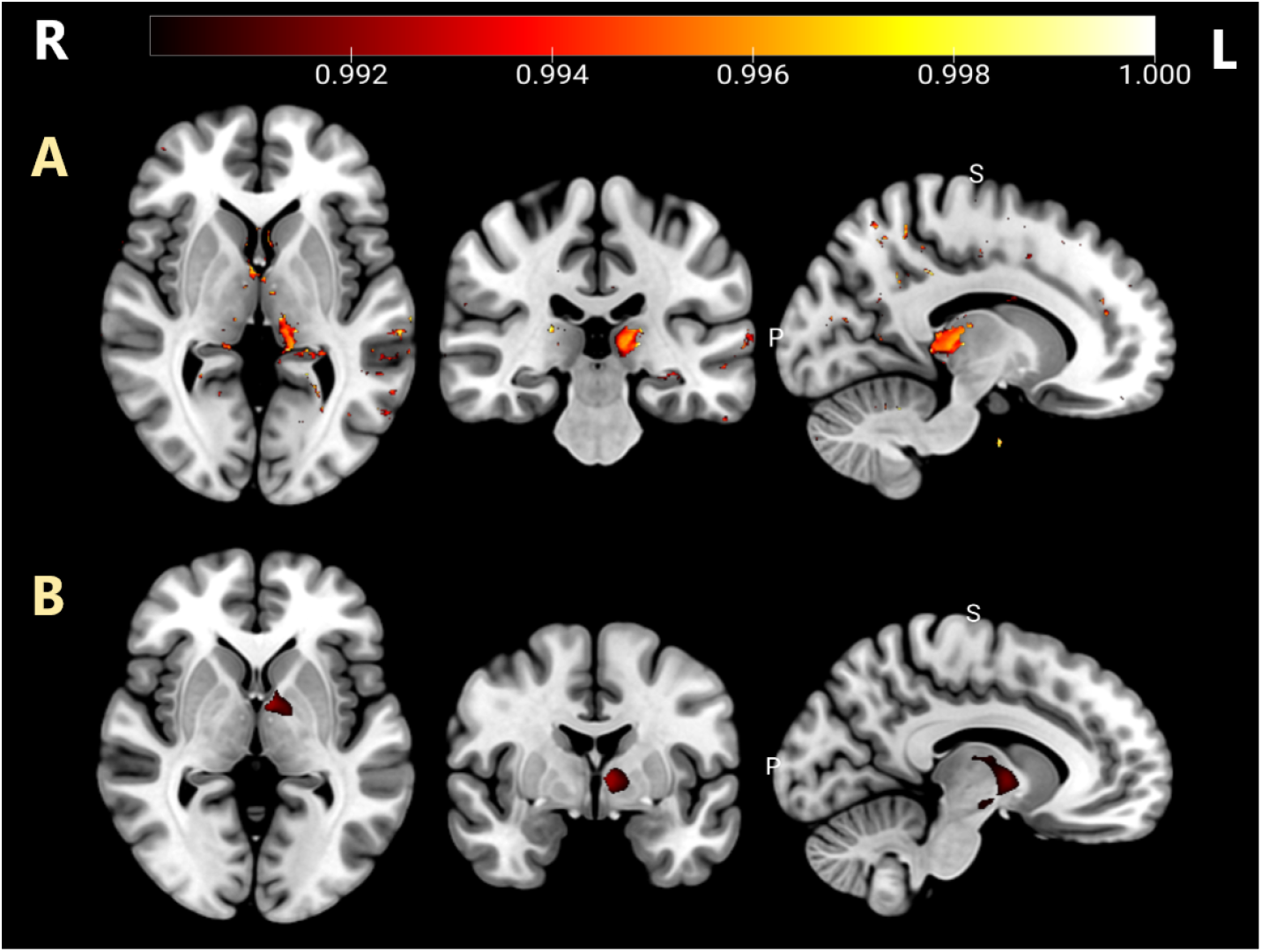
MTR clusters demonstrating regions including lower myelin (red-yellow) in NF1 compared to controls. Grey matter myelin shows clusters in (A) left thalamus extended to the left hippocampus. (B) A large extended cluster of reduced white matter myelin is seen in thalamus, pallidum and midbrain in the left hemisphere.

**Fig. 4.**
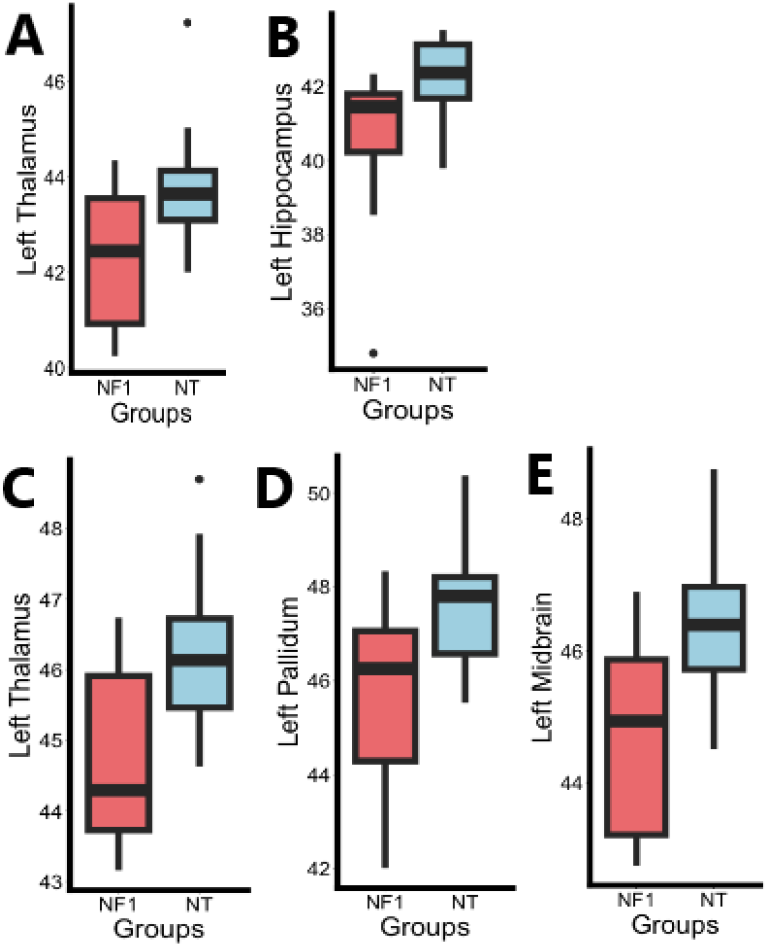
The boxplots for MTR represent the lower myelin in NF1 (red) than the controls (blue) in the grey matter including (A) left thalamus and (B) left hippocampus, and white matter including (C) left thalamus, (D) left pallidum and (E) left midbrain.

### T1W/T2W ratio study

We then sought to validate the MTR findings in a larger NF1 cohort using the routinely collected T1W and T2W structural images (NF1 =55, controls = 20).

The T1W/T2W ratio grey matter showed significantly reduced myelin in the bilateral caudate and amygdala, left posterior superior temporal gyrus, left middle temporal gyrus (temporo-occipital cortex) left accumbens and right subcallosal cortex in NF1 compared to controls (Fig. 5 and 6, Table 3). The white matter showed significantly reduced myelin in NF1 compared to controls in the bilateral cerebellum, extending to the cerebellar peduncles. No significant association between myelin (as measured by T1W/T2W ratio) was found with working memory performance (accuracy or reaction time) or with adaptive behaviour (supplementary section: 3).

**Fig. 5.**
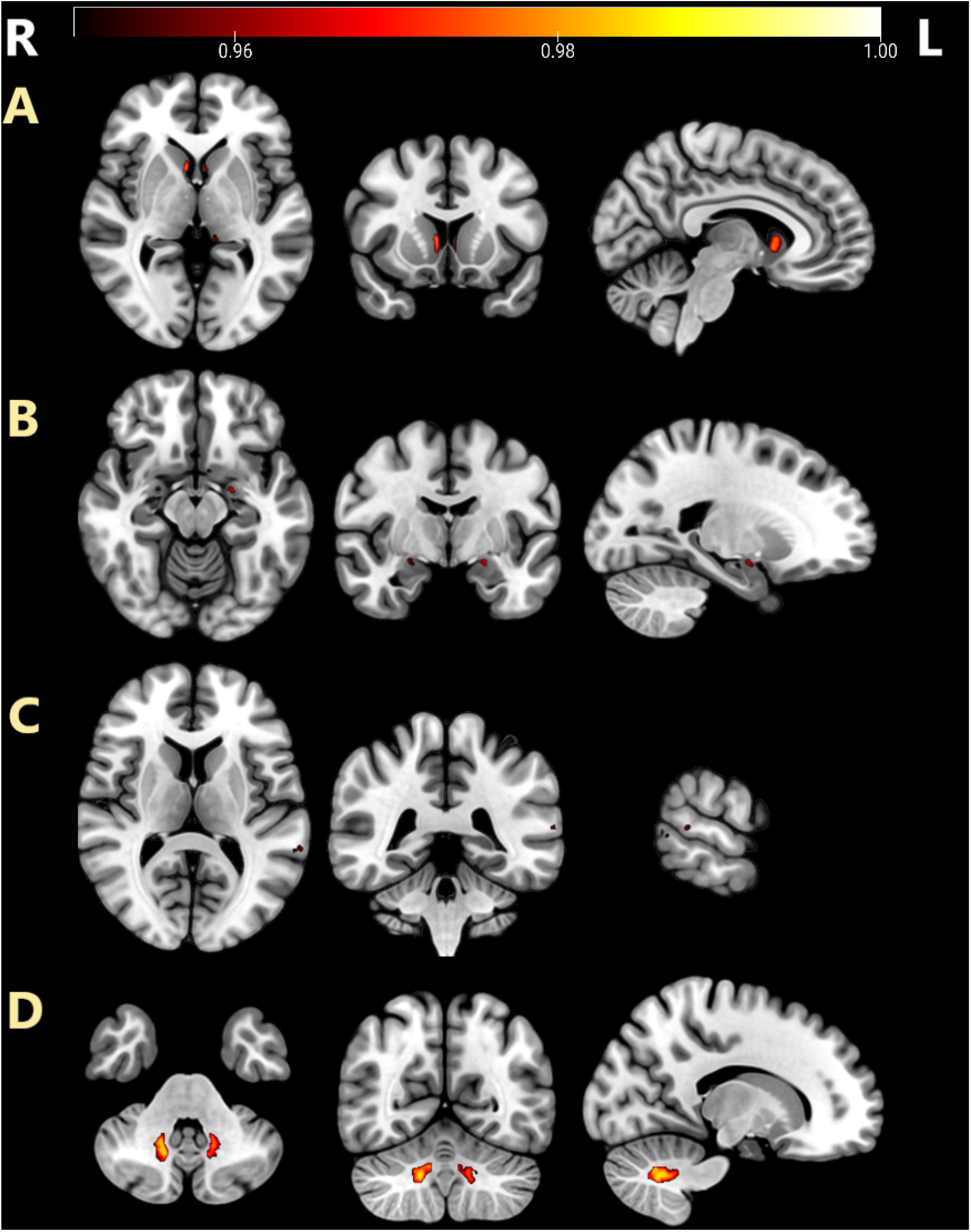
T1W/T2W ratio clusters (on MNI152 brain template overlay) demonstrating significantly reduced myelin (red-yellow) for NF1 compared to controls. Reduced-grey matter myelin is shown in (A) bilateral caudate, (B) bilateral amygdala, left thalamus, (C) left posterior superior temporal gyrus. Reduced white matter myelin is shown in (D) bilateral cerebellum and cerebellar peduncles. A threshold (p=0.05) was used to delineate all clusters from family wise error corrected maps.

**Fig. 6.**
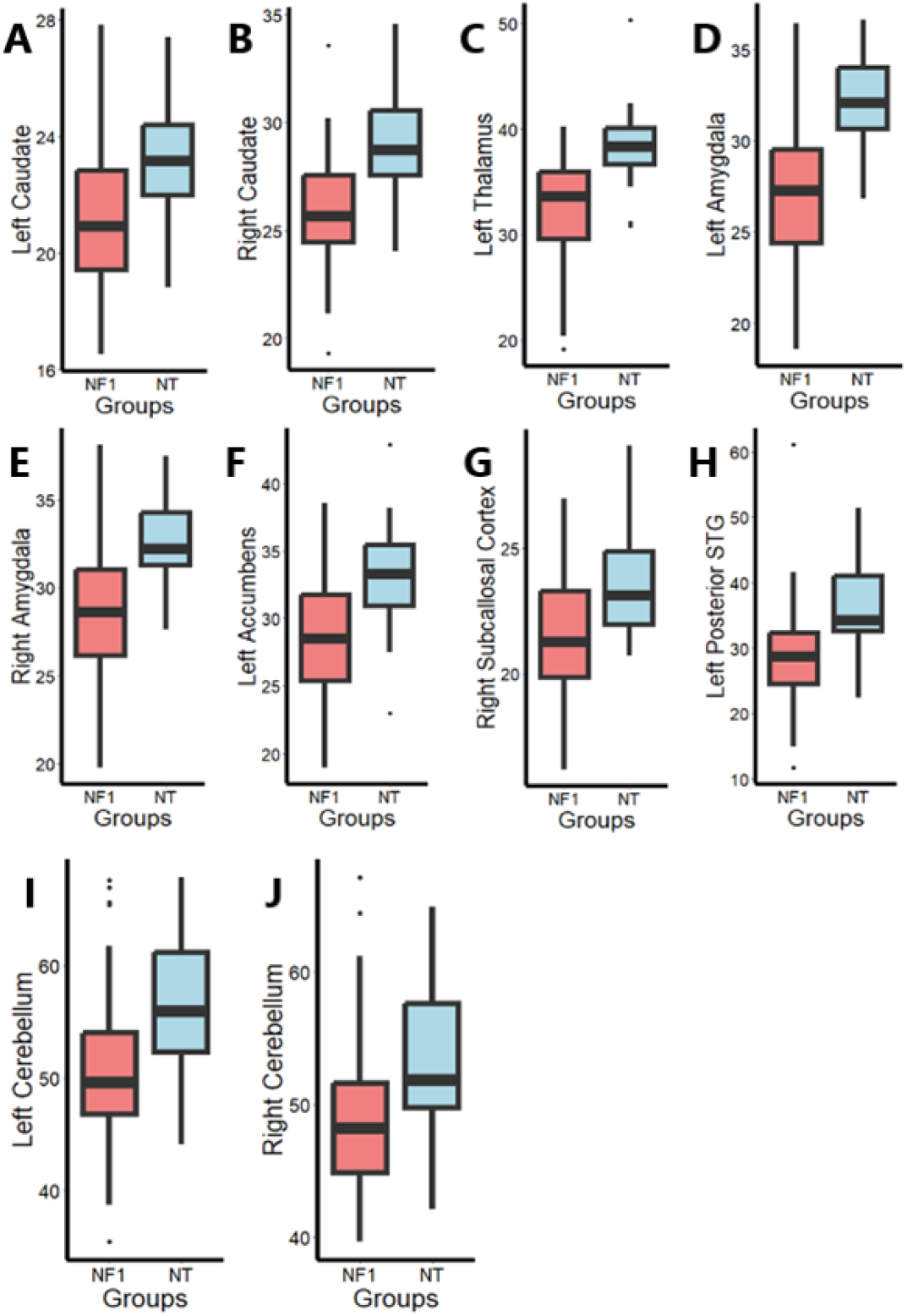
The boxplots for T1W/T2W ratio represents lower myelin in NF1 (red) than the neurotypical controls (blue) in the grey matter myelin including (A) left caudate, (B) right caudate, (C) left thalamus, (D) left amygdala, (E) right amygdala, (F) left accumbens, (G) right subcallosal cortex and (H) left posterior superior temporal gyrus and white matter myelin including (I) left cerebellum and (J) right cerebellum.

**Table 3.** Case-control comparison of T1W/T2W ratio between NF1 and controls *k-Cluster size, F-effect size, p<0.05 (Family Wise Error corrected)*

| Brain regions | Hemisphere | MNI-coordinates |  |  | k | F | p-FWE |
| --- | --- | --- | --- | --- | --- | --- | --- |
| Grey matter myelin |  | x | y | z |  |  |  |
| Caudate | Left | -5 | 11 | 1 | 194 | 3.81 | 0.035 |
| Caudate | Right | 7 | 11 | -1 | 236 | 4.14 | 0.020 |
| Thalamus | Left | -12 | -35 | 2 | 60 | 4.11 | 0.040 |
| Amygdala | Left | -22 | -7 | -14 | 71 | 4.84 | 0.035 |
| Amygdala | Right | 23 | -6 | -12 | 50 | 4.54 | 0.042 |
| Posterior STG | Left | -65 | -42 | 10 | 64 | 3.68 | 0.043 |
| Accumbens | Left | -8 | 5 | -14 | 28 | 3.97 | 0.043 |
| Subcallosal Cortex | Right | 2 | 9 | -8 | 16 | 4.38 | 0.043 |
| White matter myelin |  |  |  |  |  |  |  |
| Cerebellum | Left | -15 | -56 | -35 | 869 | 4.39 | 0.023 |
| Cerebellum | Right | 15 | -59 | -36 | 992 | 4.61 | 0.014 |

## Discussion

This study provides convergent multi-modal evidence for subcortical and cerebellar hypomyelination in a pediatric NF1 cohort. Using both MTR and T1W/T2W imaging, we demonstrated reduced myelination in thalamus, basal ganglia, and cerebellum compared to typically developing controls. These findings replicate across independent imaging modalities, establishing robust evidence for myelin deficits in NF1. However, myelin metrics did not correlate significantly with working memory or adaptive behavior measures.

Our subcortical hypomyelination findings align with preclinical evidence demonstrating oligodendrocyte dysfunction in *Nf1* animal models. (López-Juárez et al., 2017, Mayes et al., 2013, Pan et al., 2024) and diffusion studies in NF1 clinical cohorts (Ferraz-Filho et al., 2012a, Ferraz-Filho et al., 2012b). Loss-of-function mutations in the NF1 gene lead to RAS/MAPK hyperactivation, which in turn impairs oligodendrocyte precursor cell differentiation — a process that is critical for myelin production and maintaining its structure and integrity, both of which are essential for fine-tuning neural circuit dynamics and neural efficiency. Critically, oligodendrocyte-specific NF1 deletion is sufficient to produce widespread reductions in fractional anisotropy (FA), impaired inter-hemispheric connectivity and impaired motor performance in mice-phenotypes that are rescued by NOS-specific inhibition (Asleh et al., 2020). These results suggest a primary oligodendrocyte pathology drives myelin structural abnormalities and compromises neuronal circuit efficiency in NF1.

Our findings contrast with a recent quantitative T1 mapping study reporting *increased* cortical white matter myelination in NF1 (Plank et al., 2025^b^), yet align with subsequent T1W/T2W data from the same group showing reduced myelination in the nucleus accumbens (Plank et al., 2025^a^). These discrepancies likely reflect differential sensitivity of imaging contrasts: quantitative T1 may be more influenced by tissue water content and macromolecular composition, whereas MTR more specifically indexes myelin lipid-protein interactions. The convergence between MTR and T1W/T2W in our study—two mechanistically distinct contrasts—strengthens confidence that observed differences reflect genuine hypomyelination rather than methodological artifacts. Critically, our findings of reduced MTR extend beyond regions with visible T2-weighted hyperintensities (T2WMH). The sensitivity analyses including patients with T2WMH still exhibited significantly reduced MTR compared to controls. This demonstrates that myelin abnormalities in NF1 represent diffuse microstructural pathology rather than focal lesion-driven effects. Histopathological studies attribute T2WMH to myelin vacuolation and spongiform changes, suggesting our MTR findings may index early or subclinical stages of this process.

Contrary to our hypothesis, this study did not find a significant association between myelin and working memory performance in NF1. The brain-behavior relationship in NF1 needs to be reassessed with a comprehensive battery of cognitive tests including digit span, Corsi block, verbal recognition memory, visual matrix, memory (Alloway et al., 2009, Conklin et al., 2007, Raghubar et al., 2010). Despite this, it is likely that the myelin differences in the subcortical structures contribute to the cognitive phenotype observed in NF1 (Plank et al., 2025^a^). The cerebellum, for example, has a well-established role in maintaining motor coordination, often impaired in NF1. Similarly, the thalamus acts as a central processing hub essential for functions such as attention, information processing and memory. It projects to several cortical areas and receives input from the basal ganglia. Previous studies have found abnormal patterns of brain metabolites including low N-acetyaspartate (NAA)/Choline (Cho) and ratios in the thalamus irrespective of the presence of T2WMH (Wang et al., 2000). Positron Emission Tomography imaging with Fluorodeoxyglucose showed reduced glucose metabolic activity specifically in the thalamus regardless of T2WMH, indicating regional hypometabolism (Ozden et al., 2024, Reinert et al., 2019, Apostolova et al., 2015, Kaplan et al., 1997).

Our findings have implications beyond NF1. Subcortical hypomyelination, particularly affecting amygdala, overlaps with observations in idiopathic autism spectrum disorder (Gibbard et al., 2018, Barnea-Goraly et al., 2004). Myelin abnormalities have been reported across neurodevelopmental conditions including ADHD, with heterogeneous findings depending on imaging modality (Dipnall et al., 2025, Wu et al., 2017). NF1 thus provides a monogenic model for investigating how oligodendrocyte dysfunction contributes to cognitive and behavioral phenotypes shared across multiple psychiatric diagnoses. The high co-occurrence of autism (10-30%) and ADHD (50%) in NF1 further positions this condition as a valuable experimental system for understanding myelin-related mechanisms underlying these common neurodevelopmental conditions.

A surprising discovery in our study was that the pituitary depth was shallower in NF1 compared to neurotypical controls. In our analysis, the pituitary depth served as a proxy measure to account for the effects of puberty. The pituitary gland plays an important role in puberty during adolescence. Specifically, the hypothalamic-pituitary-gonadal axis stimulates the production of sex hormones, including testosterone and estradiol, in males and females, respectively, beginning from early adolescence (Genc et al., 2023). These sex hormones are believed to accelerate the maturation of brain myelin during puberty in adolescence (Herting et al., 2012). Sex hormones, which are associated with pathophysiological differences in neurodevelopmental conditions such as NF1 may affect the size of the pituitary gland during adolescence (Diggs-Andrews et al., 2014). Evidence from previous studies suggests that neurofibromin plays a critical role in hypothalamic-pituitary axis function and suboptimal pubertal growth acceleration (Zessis et al., 2018, Hegedus et al., 2008). Pituitary depth may therefore index altered hypothalamic pituitary signalling in NF1, potentially contributing to the atypical pubertal development and brain maturation observed in this condition.

MTR provides quantitative myelin assessment in both grey and white matter, with established sensitivity in demyelinating disorders, though requiring longer acquisition times compared to T1W/T2W structural imaging. T1W/T2W myelin mapping offers practical advantages, including utilising routine clinical sequences. However, the T1W/T2W mapping data must be interpreted with caution because the varying T1W and T2W signal intensities may sometimes mimic the biological properties of the tissue including iron, calcium and water concentration (Norbom et al., 2020). Validating the T1W/T2W study findings with other quantitative myelin techniques like MTR will improve reliability of this metric (Hagiwara et al., 2018, Arshad et al., 2017). Limitations of our study include the relatively small sample size for the MTR sequences and the limited range of cognitive tests used to examine the brain-behaviour relationships. Future investigations should employ comprehensive neuropsychological batteries to capture the full spectrum of executive, attentional, motor and behavioral domains affected in NF1. Longitudinal designs tracking myelin development across childhood and adolescence would elucidate developmental trajectories and potential sensitive periods for intervention.

In summary, using complementary neuroimaging modalities, we establish robust evidence for subcortical and cerebellar hypomyelination in pediatric NF1. These deficits extend beyond focal lesions, reflecting diffuse oligodendrocyte dysfunction driven by *NF1* haploinsufficiency and Ras/MAPK dysregulation. While direct myelin-cognition relationships require further investigation with expanded assessment tools, the affected subcortical structures support multiple domains impaired in the general NF1 population. Our findings of reduced pituitary depth suggest additional hormonal contributions to atypical myelin development. MTR and T1W/T2W ratio may serve as quantitative biomarkers for clinical trials targeting oligodendrocyte function or myelin repair in NF1 and related neurodevelopmental disorders. Given the transdiagnostic relevance of myelin abnormalities across autism, ADHD, and other psychiatric conditions, insights from NF1 may inform mechanistic understanding and therapeutic development for a broad range of cognitive and behavioral phenotypes.

## Data Availability

Anonymized data used in this study can be made available upon request from the authors.

## Acknowledgements

VAC was funded by the NIHR Manchester Biomedical Research Centre (NIHR203308). The views expressed are those of the author(s) and not necessarily those of the NIHR or the Department of Health and Social Care”. SG was awarded a Francis Collins Scholarship to support this study through Neurofibromatosis Therapeutic Acceleration Program (NTAP). This research was also supported by the NIHR Biomedical Research Centre, Manchester and the National Institute for Health and Care Research (NIHR) Mental Health Translational Research Collaboration, hosted by the NIHR Oxford Health Biomedical Research Centre. We thank the research radiographer’s team (Amy Watkins and colleagues) at Clinical Research Facility for their support to MRI scans and our former research assistants Georgia Birchenough and Jessica Ward for participant recruitment.

## Authorship contributions

Varun Arunachalam Chandran: Formal Analysis, Investigation, Methodology, Writing-original manuscript Caroline Lea-Carnall: Writing – Methodology, Investigation and Writing – review & editing

Anna Wild: Methodology

Matthew McCowen: Methodology

Yuping Yang: Methodology

Grace Vassallo: Investigation

Jonathan Green: Investigation

William Lloyd: Methodology, Validation, Writing – review & editing

Hamied Haroon: Methodology and Writing – review & editing

Stavros Stivaros: Investigation-Clinical evaluation

Nils Muhlert: Supervision, Methodology, Validation, Writing – review & editing

Shruti Garg: Conceptualization, Supervision, Investigation, Writing – review & editing

## Data sharing

Anonymized data used in this study can be made available upon request from the authors.

## Declaration

The authors declare that they have no known competing financial interests or personal relationships that could have appeared to influence the work reported in this paper.

## Notes

### Competing Interest Statement

The authors have declared no competing interest.

### Author Declarations

The study was initiated after receiving approval from the NHS Research Ethics Committee (reference number: 18/NW/0762).

## References

1. Alex, A. M., Buss, C., Davis, E. P., de Los Campos, G., Donald, K. A., Fair, D. A., & Knickmeyer, R. (2023). Genetic influences on the developing young brain and risk for neuropsychiatric disorders. Biological psychiatry, 93(10), 905–920.

2. Alloway, T. P., Gathercole, S. E., Kirkwood, H., & Elliott, J. (2009). The cognitive and behavioral characteristics of children with low working memory. Child development, 80(2), 606–621.

3. Aoki, S., Barkovich, A. J., Nishimura, K., Kjos, B. O., Machida, T., Cogen, P., & Norman, D. (1989). Neurofibromatosis types 1 and 2: cranial MR findings. Radiology, 172(2), 527–534.

4. Apostolova, I., Derlin, T., Salamon, J., Amthauer, H., Granström, S., Brenner, W., & Buchert, R. (2015). Cerebral glucose metabolism in adults with neurofibromatosis type 1. Brain Research, 1625, 97–101.

5. Arshad, M., Stanley, J. A., & Raz, N. (2017). Test–retest reliability and concurrent validity of in vivo myelin content indices: Myelin water fraction and calibrated T1w/T2w image ratio. Human brain mapping, 38(4), 1780–1790.

6. Asleh, J., Shofty, B., Cohen, N., Kavushansky, A., López-Juárez, A., Constantini, S., & Kahn, I. (2020). Brain-wide structural and functional disruption in mice with oligodendrocyte-specific Nf1 deletion is rescued by inhibition of nitric oxide synthase. Proceedings of the National Academy of Sciences, 117(36), 22506–22513.

7. Barnea-Goraly, N., Kwon, H., Menon, V., Eliez, S., Lotspeich, L., & Reiss, A. L. (2004). White matter structure in autism: preliminary evidence from diffusion tensor imaging. Biological psychiatry, 55(3), 323–326.

8. Baudou, E., Nemmi, F., Biotteau, M., Maziero, S., Assaiante, C., Cignetti, F., & Chaix, Y. (2020). Are morphological and structural MRI characteristics related to specific cognitive impairments in neurofibromatosis type 1 (NF1) children?. European Journal of Paediatric Neurology, 28, 89–100.

9. Bawden, H., Dooley, J., Buckley, D., Camfield, P., Gordon, K., Riding, M., & Llewellyn, G. (1996). MRI and nonverbal cognitive deficits in children with neurofibromatosis 1. Journal of Clinical and Experimental Neuropsychology, 18(6), 784–792.

10. Bennett, M. R., Rizvi, T. A., Karyala, S., McKinnon, R. D., & Ratner, N. (2003). Aberrant growth and differentiation of oligodendrocyte progenitors in neurofibromatosis type 1 mutants. Journal of Neuroscience, 23(18), 7207–7217.

11. Carter, A. S., Volkmar, F. R., Sparrow, S. S., Wang, J. J., Lord, C., Dawson, G., & Schopler, E. (1998). The Vineland Adaptive Behavior Scales: supplementary norms for individuals with autism. Journal of autism and developmental disorders, 28, 287–302.

12. Chabernaud, C., Sirinelli, D., Barbier, C., Cottier, J. P., Sembely, C., Giraudeau, B., & Castelnau, P. (2009). Thalamo-striatal T2-weighted hyperintensities (unidentified bright objects) correlate with cognitive impairments in neurofibromatosis type 1 during childhood. Developmental neuropsychology, 34(6), 736–748.

13. Chen, J. T., Collins, D. L., Freedman, M. S., Atkins, H. L., Arnold, D. L., & Canadian MS/BMT Study Group. (2005). Local magnetization transfer ratio signal inhomogeneity is related to subsequent change in MTR in lesions and normal-appearing white-matter of multiple sclerosis patients. Neuroimage, 25(4), 1272–1278.

14. Christian, I. R., Liuzzi, M. T., Yu, Q., Kryza-Lacombe, M., Monk, C. S., Jarcho, J., & Wiggins, J. L. (2022). Context-dependent amygdala-prefrontal connectivity in youths with autism spectrum disorder. Research in Autism Spectrum Disorders, 91, 101913.

15. Conklin, H. M., Luciana, M., Hooper, C. J., & Yarger, R. S. (2007). Working memory performance in typically developing children and adolescents: Behavioral evidence of protracted frontal lobe development. Developmental neuropsychology, 31(1), 103–128.

16. Corrigan, N. M., Yarnykh, V. L., Hippe, D. S., Owen, J. P., Huber, E., Zhao, T. C., & Kuhl, P. K. (2021). Myelin development in cerebral gray and white matter during adolescence and late childhood. Neuroimage, 227, 117678.

17. DiMario, F. J., & Ramsby, G. (1998). Magnetic resonance imaging lesion analysis in neurofibromatosis type 1. Archives of Neurology, 55(4), 500–505.

18. Dipnall, L. M., Fuelscher, I., Yang, J. Y., Chen, J., Craig, J. M., Anderson, V., & Silk, T. J. (2025). Brain Myelin in Children With Attention-Deficit/Hyperactivity Disorder: A Longitudinal T1-Weighted/T2-Weighted Ratio Study. Biological Psychiatry: Cognitive Neuroscience and Neuroimaging.

19. de Blank, P., Nishiyama, A., & López-Juárez, A. (2023). A new era for myelin research in Neurofibromatosis type 1. Glia, 71(12), 2701–2719.

20. Deoni, S. C., Dean III, D. C., Remer, J., Dirks, H., & O’Muircheartaigh, J. (2015). Cortical maturation and myelination in healthy toddlers and young children. Neuroimage, 115, 147–161.

21. Diggs-Andrews, K. A., Brown, J. A., Gianino, S. M., Rubin, J. B., Wozniak, D. F., & Gutmann, D. H. (2014). Sex is a major determinant of neuronal dysfunction in neurofibromatosis type 1. Annals of neurology, 75(2), 309–316.

22. DiPaolo, D. P., Zimmerman, R. A., Rorke, L. B., Zackai, E. H., Bilaniuk, L. T., & Yachnis, A. T. (1995). Neurofibromatosis type 1: pathologic substrate of high-signal-intensity foci in the brain. Radiology, 195(3), 721–724.

23. Douet, V., Chang, L., Cloak, C., & Ernst, T. (2014). Genetic influences on brain developmental trajectories on neuroimaging studies: from infancy to young adulthood. Brain imaging and behavior, 8, 234–250.

24. Feldmann, R., Denecke, J., Grenzebach, M., Schuierer, G., & Weglage, J. (2003). Neurofibromatosis type 1: motor and cognitive function and T2-weighted MRI hyperintensities. Neurology, 61(12), 1725–1728.

25. Ferraz-Filho, J. R., da Rocha, A. J., Muniz, M. P., Souza, A. S., Goloni-Bertollo, E. M., & Pavarino-Bertelli, É.C. (2012). Diffusion tensor MR imaging in neurofibromatosis type 1: expanding the knowledge of microstructural brain abnormalities. Pediatric radiology, 42, 449–454.

26. Ferraz-Filho, J. R. L., da Rocha, A. J., Muniz, M. P., Souza, A. S., Goloni-Bertollo, E. M., & Pavarino-Bertelli, É.C. (2012). Unidentified bright objects in neurofibromatosis type 1: conventional MRI in the follow-up and correlation of microstructural lesions on diffusion tensor images. European journal of paediatric neurology, 16(1), 42–47.

27. Genc, S., Raven, E. P., Drakesmith, M., Blakemore, S. J., & Jones, D. K. (2023). Novel insights into axon diameter and myelin content in late childhood and adolescence. Cerebral Cortex, 33(10), 6435–6448.

28. Gibbard, C. R., Ren, J., Skuse, D. H., Clayden, J. D., & Clark, C. A. (2018). Structural connectivity of the amygdala in young adults with autism spectrum disorder. Human brain mapping, 39(3), 1270–1282.

29. Glasser, M. F., & Van Essen, D. C. (2011). Mapping human cortical areas in vivo based on myelin content as revealed by T1-and T2-weighted MRI. Journal of neuroscience, 31(32), 11597–11616.

30. Goh, W. H., Khong, P. L., Leung, C. S., & Wong, V. C. (2004). T 2-weighted hyperintensities (unidentified bright objects) in children with neurofibromatosis 1: their impact on cognitive function. Journal of child neurology, 19(11), 853–858.

31. Greenwood, R. S., Tupler, L. A., Whitt, J. K., Buu, A., Dombeck, C. B., Harp, A. G., & MacFall, J. R. (2005). Brain morphometry, T2-weighted hyperintensities, and IQ in children with neurofibromatosis type 1. Archives of neurology, 62(12), 1904–1908.

32. Hachon, C., Iannuzzi, S., & Chaix, Y. (2011). Behavioural and cognitive phenotypes in children with neurofibromatosis type 1 (NF1): the link with the neurobiological level. Brain and Development, 33(1), 52–61.

33. Hagiwara, A., Hori, M., Kamagata, K., Warntjes, M., Matsuyoshi, D., Nakazawa, M., & Aoki, S. (2018). Myelin measurement: comparison between simultaneous tissue relaxometry, magnetization transfer saturation index, and T1w/T2w ratio methods. Scientific reports, 8(1), 10554

34. Han, Yong, Hangzhou Wang, and Yulun Huang. “T2 Hyperintensities in Children with Neurofibromatosis Type 1.” World Neurosurgery 192 (2024): e480–e485.

35. Haroutunian, V., Katsel, P., Roussos, P., Davis, K. L., Altshuler, L. L., & Bartzokis, G. (2014). Myelination, oligodendrocytes, and serious mental illness. Glia, 62(11), 1856–1877.

36. Hegedus, B., Yeh, T. H., Lee, D. Y., Emnett, R. J., Li, J., & Gutmann, D. H. (2008). Neurofibromin regulates somatic growth through the hypothalamic–pituitary axis. Human molecular genetics, 17(19), 2956–2966.

37. Henkelman, R. M., Stanisz, G. J., & Graham, S. J. (2001). Magnetization transfer in MRI: a review. NMR in Biomedicine: An International Journal Devoted to the Development and Application of Magnetic Resonance In Vivo, 14(2), 57–64.

38. Herting, M. M., Maxwell, E. C., Irvine, C., & Nagel, B. J. (2012). The impact of sex, puberty, and hormones on white matter microstructure in adolescents. Cerebral cortex, 22(9), 1979–1992.

39. Huijbregts, S., Jahja, R., De Sonneville, L. E. O., De Breij, S., & SWAAB-BARNEVELD, H. A. N. N. A. (2010). Social information processing in children and adolescents with neurofibromatosis type 1. Developmental Medicine & Child Neurology, 52(7), 620–625.

40. Hyman, S. L., Gill, D. S., Shores, E. A., Steinberg, A., Joy, P., Gibikote, S. V., & North, K. N. (2003). Natural history of cognitive deficits and their relationship to MRI T2-hyperintensities in NF1. Neurology, 60(7), 1139–1145.

41. Jakovcevski, I., Filipovic, R., Mo, Z., Rakic, S., & Zecevic, N. (2009). Oligodendrocyte development and the onset of myelination in the human fetal brain. Frontiers in neuroanatomy, 3, 665.

42. Jenkinson, M., & Smith, S. (2001). A global optimisation method for robust affine registration of brain images. Medical image analysis, 5(2), 143–156.

43. Kaplan, A. M., Chen, K., Lawson, M. A., Wodrich, D. L., Bonstelle, C. T., & Reiman, E. M. (1997). Positron emission tomography in children with neurofibromatosis-1. Journal of Child Neurology, 12(8), 499–506.

44. Karlsgodt, K. H., Rosser, T., Lutkenhoff, E. S., Cannon, T. D., Silva, A., & Bearden, C. E. (2012). Alterations in white matter microstructure in neurofibromatosis-1.

45. Koini, M., Rombouts, S. A. R. B., Veer, I. M., Van Buchem, M. A., & Huijbregts, S. C. J. (2017). White matter microstructure of patients with neurofibromatosis type 1 and its relation to inhibitory control. Brain imaging and behavior, 11(6), 1731–1740.

46. Lee, T. S. J., Chopra, M., Kim, R. H., Parkin, P. C., & Barnett-Tapia, C. (2023). Incidence and prevalence of neurofibromatosis type 1 and 2: a systematic review and meta-analysis. Orphanet Journal of Rare Diseases, 18(1), 292.

47. Legius, E., Messiaen, L., Wolkenstein, P., Pancza, P., Avery, R. A., Berman, Y., & Plotkin, S. R. (2021). Revised diagnostic criteria for neurofibromatosis type 1 and Legius syndrome: an international consensus recommendation. Genetics in Medicine, 23(8), 1506–1513.

48. Lin, L., Chen, Y., Dai, Y., Yan, Z., Zou, M., Zhou, Q., & Su, S. (2024). Quantification of myelination in children with attention-deficit/hyperactivity disorder: a comparative assessment with synthetic MRI and DTI. European Child & Adolescent Psychiatry, 33(6), 1935–1944.

49. Mayes, D. A., Rizvi, T. A., Titus-Mitchell, H., Oberst, R., Ciraolo, G. M., Vorhees, C. V., & Ratner, N. (2013). Nf1 loss and Ras hyperactivation in oligodendrocytes induce NOS-driven defects in myelin and vasculature. Cell reports, 4(6), 1197–1212.

50. Moccia, M., van de Pavert, S., Eshaghi, A., Haider, L., Pichat, J., Yiannakas, M., & Ciccarelli, O. (2020). Pathologic correlates of the magnetization transfer ratio in multiple sclerosis. Neurology, 95(22), e2965–e2976.

51. Moore III, B. D., Slopis, J. M., Schomer, D., Jackson, E. F., & Levy, B. M. (1996). Neuropsychological significance of areas of high signal intensity on brain MRIs of children with neurofibromatosis. Neurology, 46(6), 1660–1668.

52. Nave, K. A., & Ehrenreich, H. (2014). Myelination and oligodendrocyte functions in psychiatric diseases. JAMA psychiatry, 71(5), 582–584.

53. Nemmi, F., Cignetti, F., Assaiante, C., Maziero, S., Audic, F., Péran, P., & Chaix, Y. (2019). Discriminating between neurofibromatosis-1 and typically developing children by means of multimodal MRI and multivariate analyses. Human Brain Mapping, 40(12), 3508–3521.

54. Nicita, F., Di Biasi, C., Sollaku, S., Cecchini, S., Salpietro, V., Pittalis, A., & Spalice, A. (2014). Evaluation of the basal ganglia in neurofibromatosis type 1. Child’s Nervous System, 30(2), 319–325.

55. Norbom, L. B., Rokicki, J., Alnæs, D., Kaufmann, T., Doan, N. T., Andreassen, O. A., & Tamnes, C. K. (2020). Maturation of cortical microstructure and cognitive development in childhood and adolescence: a T1w/T2w ratio MRI study. Human brain mapping, 41(16), 4676–4690.

56. O’Muircheartaigh, J., Dean III, D. C., Ginestet, C. E., Walker, L., Waskiewicz, N., Lehman, K., & Deoni, S. C. (2014). White matter development and early cognition in babies and toddlers. Human brain mapping, 35(9), 4475–4487.

57. Özden, C., Mautner, V. F., Farschtschi, S., Molwitz, I., Ristow, I., Bannas, P., & Buchert, R. (2024). Asymmetry of thalamic hypometabolism on FDG-PET/CT in neurofibromatosis type 1: Association with peripheral tumor burden. Journal of Neuroimaging, 34(1), 138–144.

58. Pan, Y., Hysinger, J. D., Yalçın, B., Lennon, J. J., Byun, Y. G., Raghavan, P., & Monje, M. (2024). Nf1 mutation disrupts activity-dependent oligodendroglial plasticity and motor learning in mice. Nature neuroscience, 27(8), 1555–1564.

59. Pardej, S. K., Glad, D. M., Casnar, C. L., Janke, K. M., & Klein-Tasman, B. P. (2022). Longitudinal investigation of early motor development in neurofibromatosis type 1. Journal of pediatric psychology, 47(2), 180–188.

60. Pareto, D., Garcia-Vidal, A., Alberich, M., Auger, C., Montalban, X., Tintoré, M., & Rovira, À. (2020). Ratio of T1-weighted to T2-weighted signal intensity as a measure of tissue integrity: comparison with magnetization transfer ratio in patients with multiple sclerosis. American Journal of Neuroradiology, 41(3), 461–463.

61. Payne, J. M., Pickering, T., Porter, M., Oates, E. C., Walia, N., Prelog, K., & North, K. N. (2014). Longitudinal assessment of cognition and T2-hyperintensities in NF1: An 18-year study. American journal of medical genetics Part A, 164(3), 661–665.

62. Piredda, G. F., Hilbert, T., Thiran, J. P., & Kober, T. (2021). Probing myelin content of the human brain with MRI: A review. Magnetic resonance in medicine, 85(2), 627–652.

63. Piscitelli, O., Digilio, M. C., Capolino, R., Longo, D., & Di Ciommo, V. (2012). Neurofibromatosis type 1 and cerebellar T2-hyperintensities: The relationship to cognitive functioning. Developmental medicine and child neurology, 54(1), 49.

64. Plank, J. R., Pardej, S. K., Raman, M. M., McNab, J., & Green, T. (2025). T1w/T2w ratio suggests reduced intracortical myelin content in youth with RASopathies. medRxiv, 2025-09.

65. Plank, J. R., Gozdas, E., Bruno, J., McGhee, C. A., Wu, H., Raman, M. M., & Green, T. (2025). Quantitative T1 mapping indicates elevated white matter myelin in children with RASopathies. Biological Psychiatry.

66. Raghubar, Kimberly P., Marcia A. Barnes, and Steven A. Hecht. “Working memory and mathematics: A review of developmental, individual difference, and cognitive approaches.” Learning and individual differences 20.2 (2010): 110–122.

67. Reinert, C. P., Schuhmann, M. U., Bender, B., Gugel, I., la Fougère, C., Schäfer, J., & Gatidis, S. (2019). Comprehensive anatomical and functional imaging in patients with type I neurofibromatosis using simultaneous FDG-PET/MRI. European journal of nuclear medicine and molecular imaging, 46(3), 776–787.

68. Remaud, J., Besnard, J., Barbarot, S., & Roy, A. (2024). Perception and recognition of primary and secondary emotions by children with neurofibromatosis type 1. Child Neuropsychology, 30(1), 188–201.

69. Salman, M. S., Hossain, S., Gorun, S., Alqublan, L., Bunge, M., & Rozovsky, K. (2018). Cerebellar radiological abnormalities in children with neurofibromatosis type 1: part 2-a neuroimaging natural history study with clinical correlations. Cerebellum & Ataxias, 5, 1–13.

70. Sandrone, S., Aiello, M., Cavaliere, C., Thiebaut de Schotten, M., Reimann, K., Troakes, C., & Dell’Acqua, F. (2023). Mapping myelin in white matter with T1-weighted/T2-weighted maps: discrepancy with histology and other myelin MRI measures. Brain Structure and Function, 228(2), 525–535.

71. Sari, S., Sari, E., Akgun, V., Ozcan, E., Ince, S., Saldir, M., & Yesilkaya, E. (2014). Measures of pituitary gland and stalk: from neonate to adolescence. Journal of Pediatric Endocrinology and Metabolism, 27(11-12), 1071-1076.

72. Siqueiros-Sanchez, M., Dai, E., McGhee, C. A., McNab, J. A., Raman, M. M., & Green, T. (2024). Impact of pathogenic variants of the Ras–mitogen-activated protein kinase pathway on major white matter tracts in the human brain. Brain Communications, 6(4), fcae274.

73. Uddin, M. N., Figley, T. D., Solar, K. G., Shatil, A. S., & Figley, C. R. (2019). Comparisons between multi-component myelin water fraction, T1w/T2w ratio, and diffusion tensor imaging measures in healthy human brain structures. Scientific reports, 9(1), 2500.

74. Wang, P. Y., Kaufmann, W. E., Koth, C. W., Denckla, M. B., & Barker, P. B. (2000). Thalamic involvement in neurofibromatosis type 1: evaluation with proton magnetic resonance spectroscopic imaging. Annals of Neurology: Official Journal of the American Neurological Association and the Child Neurology Society, 47(4), 477–484.

75. West, K. L., Kelm, N. D., Carson, R. P., Gochberg, D. F., Ess, K. C., & Does, M. D. (2018). Myelin volume fraction imaging with MRI. Neuroimage, 182, 511–521.

76. Wolff, S. D., & Balaban, R. S. (1989). Magnetization transfer contrast (MTC) and tissue water proton relaxation in vivo. Magnetic resonance in medicine, 10(1), 135–144.

77. Wu, Z. M., Bralten, J., Cao, Q. J., Hoogman, M., Zwiers, M. P., An, L., & Wang, Y. F. (2017). White matter microstructural alterations in children with ADHD: categorical and dimensional perspectives. Neuropsychopharmacology, 42(2), 572–580.

78. Yeung, M. S., Zdunek, S., Bergmann, O., Bernard, S., Salehpour, M., Alkass, K., & Frisén, J. (2014). Dynamics of oligodendrocyte generation and myelination in the human brain. Cell, 159(4), 766–774.

79. Zessis, N. R., Gao, F., Vadlamudi, G., Gutmann, D. H., & Hollander, A. S. (2018). Height growth impairment in children with neurofibromatosis type 1 is characterized by decreased pubertal growth velocity in both sexes. Journal of child neurology, 33(12), 762–766.

80. Zhang, S., Jiang, L., Hu, Z., Liu, W., Yu, H., Chu, Y., & Chen, Y. (2024). T1w/T2w ratio maps identify children with autism spectrum disorder and the relationships between myelin-related changes and symptoms. Progress in Neuro-Psychopharmacology and Biological Psychiatry, 111040.

