## Supplementary Materials for "Mapping myelin alterations in Neurofibromatosis Type 1 using Magnetization Transfer and T1W/T2W Ratio Imaging"

**Supplementary section: 1**

| Factor variables | Exploratory study sample | | Validation study sample | |
| --- | --- | --- | --- | --- |
|  | Factor-1 | Factor-2 | Factor-1 | Factor-2 |
| Visuospatial 0-back accuracy | 0.658 | - | - | 0.640 |
| Visuospatial 1-back accuracy | 0.782 | - | - | 0.766 |
| Visuospatial 2-back accuracy | 0.905 | - | - | 0.897 |
| Visuospatial 3-back accuracy | 0.749 | - | - | 0.766 |
| Visuospatial 0-back RT | -0.560 | **0.452** | 0.667 | - |
| Visuospatial 1-back RT | - | 0.856 | 0.891 | - |
| Visuospatial 2-back RT | - | 0.854 | 0.805 | - |
| Visuospatial 3-back RT | - | 0.894 | 0.838 | - |

**Table 5:** Factor loadings related to visuospatial working memory

**Supplementary section: 2**

| **Brain regions** | **Hemisphere** | **MNI-coordinates** | | | **k** | **F** | **p (uncorrected)** |
| --- | --- | --- | --- | --- | --- | --- | --- |
| **GM myelin (Accuracy)** |  | **x** | **y** | **z** |  |  |  |
| Middle Frontal Gyrus | Left | -36 | 4 | 34 | 43 | 4.17 | 0.001 |
| Frontal Orbital Cortex | Left | -39 | 30 | -1 | 42 | 4.29 | 0.001 |
| Lateral Occipital Cortex | Left | -35 | -72 | 52 | 39 | 5.23 | 0.001 |
| **GM myelin (Reaction Time)** |  |  |  |  |  |  |  |
| Posterior STS | Right | 37 | -46 | 10 | 45 | 3.35 | 0.001 |
| Postcentral Gyrus | Left | -57 | -16 | 47 | 42 | 5.2 | 0.001 |
| Frontal Orbital Cortex | Right | 14 | 21 | -16 | 37 | 4.51 | 0.001 |
| Middle Temporal Gyrus | Right | 61 | -12 | -24 | 35 | 4.45 | 0.001 |
| Inferior Frontal Gyrus | Right | 42 | 9 | 18 | 34 | 5.11 | 0.001 |
| Temporal Pole | Left | -39 | 16 | -25 | 34 | 3.8 | 0.001 |
| Supramarginal Gyrus | Left | -60 | -38 | 39 | 32 | 4.86 | 0.001 |
| Precentral Gyrus | Right | 18 | -11 | 54 | 30 | 5.07 | 0.001 |

**Table 6:** Positive association between MTR grey matter myelin and working memory performance in NF1 and controls

*GM-Grey Matter, WM-White Matter, k-Cluster size, F-effect size*

| **Brain regions** | **Hemisphere** | **MNI-coordinates** | | | **k** | **F** | **p (uncorrected)** |
| --- | --- | --- | --- | --- | --- | --- | --- |
| **WM myelin (Accuracy)** |  | **x** | **y** | **z** |  |  |  |
| Middle Frontal Gyrus | Left | -34 | 18 | 32 | 106 | 5.35 | 0.001 |
| Supple Motor Cortex | Right | 11 | -6 | 56 | 42 | 4.76 | 0.001 |
| Occipital Fusiform Gyrus | Right | 38 | -68 | -9 | 33 | 4.71 | 0.001 |
| **WM myelin (Reaction Time)** |  |  |  |  |  |  |  |
| Frontal Orbital Cortex | Right | 27 | 27 | -8 | 118 | 3.3 | 0.001 |
| Paracingulate Gyrus | Right | 2 | 46 | 0 | 61 | 3.35 | 0.001 |
| Postcentral Gyrus | Left | -38 | -18 | 41 | 58 | 3.35 | 0.001 |
| Insular Cortex | Left | -39 | -21 | -1 | 57 | 3.35 | 0.001 |
| Caudate | Right | 10 | 13 | 11 | 53 | 3.35 | 0.001 |
| Insular Cortex | Right | 39 | -20 | 2 | 52 | 3.35 | 0.001 |
| Cerebellum | Left | -29 | -56 | -33 | 51 | 3.35 | 0.001 |
| Pons | Left | -6 | -15 | -32 | 45 | 3.61 | 0.001 |
| Middle Temporal Gyrus | Left | -58 | -13 | -27 | 40 | 4.71 | 0.001 |
| Cerebellum | Right | 31 | -73 | -33 | 39 | 3.75 | 0.001 |
| Lateral Occipital Cortex | Right | 50 | -73 | -9 | 37 | 5.48 | 0.001 |
| Precentral Gyrus | Left | -55 | 1 | 17 | 34 | 4.85 | 0.001 |
| Precentral Gyrus | Right | 19 | -28 | 60 | 31 | 4.77 | 0.001 |

**Table 7:** Positive association between MTR white matter myelin and working memory performance in NF1 and controls

| **Brain regions** | **Hemisphere** | **MNI-coordinates** | | | **k** | **F** | **p (uncorrected)** |
| --- | --- | --- | --- | --- | --- | --- | --- |
| **GM myelin (Accuracy)** |  | **x** | **y** | **z** |  |  |  |
| Precentral Gyrus | Right | 23 | -14 | 70 | 88 | 4.57 | 0.001 |
| Middle Frontal Gyrus | Left | -32 | 20 | 55 | 81 | 4.01 | 0.001 |
| Postcentral Gyrus | Left | -21 | -37 | 69 | 66 | 4.56 | 0.001 |
| Parahippocampal Gyrus | Left | -19 | 1 | -33 | 61 | 4.98 | 0.001 |
| Angular Gyrus | Right | 47 | -48 | 50 | 51 | 5.32 | 0.001 |
| **GM myelin (Reaction Time)** |  |  |  |  |  |  |  |
| Cerebellum | Right | 34 | -66 | -40 | 306 | 4.21 | 0.001 |
| Posterior STS | Right | 37 | -45 | 10 | 83 | 5.75 | 0.001 |
| Lateral Occipital Cortex | Right | 47 | -73 | -9 | 65 | 4.51 | 0.001 |
| Posterior Cingulate Gyrus | Right | 3 | -31 | 45 | 63 | 4.14 | 0.001 |
| Temporal Pole | Left | -42 | 18 | -25 | 62 | 4.52 | 0.001 |
| Cerebellum | Left | -16 | -69 | -54 | 61 | 4.59 | 0.001 |
| Occipital pole | Left | -1 | -98 | -17 | 60 | 6.77 | 0.001 |

**Table 8**: Positive association between T1W/T2W grey matter myelin and working memory performance in NF1 and controls

| **Brain regions** | **Hemisphere** | **MNI-coordinates** | | | **k** | **F** | **p (uncorrected)** |
| --- | --- | --- | --- | --- | --- | --- | --- |
| **WM myelin (Accuracy)** |  | **x** | **y** | **z** |  |  |  |
| Supramarginal Gyrus | Right | 55 | -29 | 26 | 213 | 5.05 | 0.001 |
| Precentral Gyrus | Right | 22 | -15 | 66 | 105 | 5.04 | 0.001 |
| Superior Temporal Gyrus | Left | -62 | -5 | 1 | 86 | 6.56 | 0.001 |
| Inferior Temporal Gyrus | Left | -66 | -34 | -23 | 67 | 4.81 | 0.001 |
| Middle Frontal Gyrus | Left | -50 | 11 | 38 | 64 | 4.81 | 0.001 |
| Middle Temporal Gyrus | Left | -64 | -9 | -21 | 63 | 5.84 | 0.001 |
| **WM myelin (Reaction Time)** |  |  |  |  |  |  |  |
| Inferior Temporal gyrus | Left | -59 | -12 | -33 | 145 | 5.75 | 0.001 |
| Paracingulate Gyrus | Right | 1 | 46 | 0 | 113 | 7.52 | 0.001 |
| Heschl’s Gyrus | Right | 40 | -19 | 1 | 88 | 5.75 | 0.001 |
| Frontal Orbital Cortex | Left | -27 | 10 | -19 | 87 | 7.06 | 0.001 |
| Insular Cortex | Right | 42 | -8 | 7 | 59 | 5.75 | 0.001 |
| Frontal Orbital Cortex | Right | 33 | 27 | -23 | 50 | 7.69 | 0.001 |

**Table 9**: Positive association between T1W/T2W white matter myelin and working memory performance in NF1 and controls

**Supplementary section: 3**

| **Brain regions** | **Hemisphere** | **MNI-coordinates** | | | **k** | **F** | **p (uncorrected)** |
| --- | --- | --- | --- | --- | --- | --- | --- |
| **GM myelin** |  | **x** | **y** | **z** |  |  |  |
| Temporal Pole | Right | 51 | 10 | -36 | 53 | 3.34 | 0.010 |
| Insular Cortex | Left | -36 | -7 | -11 | 45 | 3.66 | 0.010 |
| Lingual Gyrus | Right | 2 | -84 | -21 | 38 | 4.65 | 0.010 |
| Precentral Gyrus | Right | 54 | 3 | 20 | 33 | 4.19 | 0.010 |
| Precuneus Cortex | Left | -8 | -62 | 32 | 33 | 3.47 | 0.010 |
| Planum Temporale | Left | -57 | -38 | 18 | 31 | 4.04 | 0.010 |
| **WM myelin** |  |  |  |  |  |  |  |
| Precentral Gyrus | Right | 55 | 8 | 12 | 116 | 3.91 | 0.010 |
| Middle Temporal Gyrus | Right | 54 | -8 | -16 | 98 | 4.34 | 0.010 |
| Pons | Left | -5 | -23 | -42 | 98 | 3.48 | 0.010 |
| Supramarginal Gyrus | Left | -53 | -31 | 43 | 45 | 4.19 | 0.010 |
| Paracingulate Gyrus | Right | 9 | 54 | -5 | 45 | 3.35 | 0.010 |
| Inferior Temporal Gyrus | Right | 46 | -25 | -28 | 41 | 3.3 | 0.010 |
| Middle Frontal Gyrus | Left | -34 | 33 | 35 | 41 | 4.19 | 0.010 |
| Superior Frontal Gyrus | Left | -4 | 38 | 39 | 41 | 4.23 | 0.010 |
| Precuneus Cortex | Left | -13 | -54 | 11 | 40 | 4.27 | 0.010 |
| Precentral Gyrus | Left | -17 | -21 | 71 | 39 | 3.76 | 0.010 |
| Supple Motor Cortex | Left | -6 | 4 | 51 | 37 | 4.27 | 0.010 |
| Lateral Occipital Cortex | Left | -21 | -64 | 45 | 37 | 4.45 | 0.010 |
| Lingual Gyrus | Right | 22 | -57 | -6 | 35 | 3.54 | 0.010 |
| Anterior Cingulate Gyrus | Left | -5 | 11 | 32 | 34 | 3.03 | 0.010 |
| Frontal Pole | Left | -25 | 49 | 23 | 30 | 3.26 | 0.010 |

**Table 10**: Positive association between MTR grey and white matter myelin and adaptive behaviour in NF1

| **Brain regions** | **Hemisphere** | **MNI-coordinates** | | | **k** | **F** | **p (uncorrected)** |
| --- | --- | --- | --- | --- | --- | --- | --- |
| **GM myelin** |  | **x** | **y** | **z** |  |  |  |
| Middle Temporal Gyrus | Left | -64 | -6 | -17 | 249 | 5.09 | 0.010 |
| Paracingulate Gyrus | Right | 2 | 43 | -10 | 160 | 4.86 | 0.010 |
| Inferior Temporal Gyrus | Right | 59 | -52 | -25 | 151 | 4.99 | 0.010 |
| Cerebellum | Left | -40 | -48 | -56 | 104 | 4.82 | 0.010 |
| **WM myelin** |  |  |  |  |  |  |  |
| Middle Temporal Gyrus | Left | -58 | -3 | -21 | 121 | 5.34 | 0.010 |

**Table 11**: Positive association between T1W/T2W grey and white matter myelin and adaptive behaviour in NF1
